# Mortality and diagnostic practice variation in interstitial lung disease admissions: insights from a multicentre UK cohort study

**DOI:** 10.1101/2025.08.01.25332724

**Authors:** Laura White, Jonathon Shaw, Bethan Powell, Nyan May Kyi, Alicia Sou, Gareth Hughes, Dilanka Tilakaratne, Conal Hayton, Trishala Raj, Vi Truong, Nashwah Ismail, Nawat Khanijoun, Rebecca Huang, Emma Hardy, Mahzaib Babar, Naayaab Khan, Martin Regan, Oby Okpala, Ragavilasini Suresh, Jerome McIntosh, Amsal Amjad, Mahum Sohail, Zainab Aslam, Amy Gadoud, Timothy Gatheral, Georges NgManKwong

## Abstract

**Background:** Interstitial lung diseases (ILD) are a heterogenous group of often progressive, unpredictable diseases. They frequently result in hospitalisations secondary to respiratory decompensation, termed ILD-related admissions. A proportion of these are due to acute exacerbations (AEILD). All are associated with high mortality but poorly characterised in real-world populations.

**Aim:** To evaluate mortality outcomes and associated risk factors following ILD-related hospital admissions, including AEILD.

**Methods:** We conducted a multicentre retrospective cohort study of primary ICD10 coded admissions for ILD between 01.01.2017 and 31.12.2019 across 11 NHS hospitals in the North West of England. AEILD events were classified using clinical criteria: a <30-day respiratory deterioration not secondary to cardiac failure, pulmonary embolism or pneumothorax. The AEILD sub-group was subsequently divided into those with CT confirmation (definite AEILD) and without CT confirmation (suspected AEILD). Primary outcome was time from admission to death. Statistical analyses included Kaplan-Meier survival and multivariate cox proportional hazards modelling.

**Results:** Of 938 admissions ILD-related admissions, 54.5% met study AEILD criteria. Overall, 90-day all-cause mortality was 40.2%. Median survival of the AEILD cohort was 107 days (95% CI 87.0 – 141.0 days) and other ILD-related admission cohort 241.0 days (95% CI 208.0 – 308.0 days), with a statistically significant difference in survival (p <0.0001). 37.6% (192/511) of AEILD events had CT confirmation. Within the AEILD sub-group, median survival was higher in the CT group (144 days vs. 100 days, p = 0.027). AEILD was independently associated with mortality in a multivariate model, and pre-admission oxygen, age and neutrophilia were associated with mortality in both ILD-admission and AEILD 90-day all-cause mortality models. Only 13.9% of admissions had documented palliative care input.

**Conclusion:** Mortality associated with ILD-related admissions is high, with AEILD events independently associated with high mortality. Findings highlight the need for improved education, improved access to palliative care and targeted AEILD research.

**Key Messages:** *What is already known on this topic.:* Hospital admissions in interstitial lung disease (ILD) carry a high risk of mortality, particularly when precipitated by an acute exacerbation (AEILD). Prior international surveys have highlighted clinician heterogeneity in the approach to AEILD, but there is very limited real-world data describing admission outcomes, diagnostic and treatment patterns from the UK.

*What this study adds.:* This study adds to the understanding that AEILD conveys poor survival outcomes and highlights age, pre-admission oxygen use and neutrophilia as poor prognostic indicators. It highlights underuse of CT for diagnostic confirmation and demonstrates that a lack of CT confirmation is associated with shorter survival in simple modelling. It also demonstrates low palliative care inpatient service utilisation.

*How this study might affect research, practice or policy.:* These findings highlight the urgent need for consistent diagnostic pathways, equitable access to CT imaging and early multidisciplinary input for AEILD. Improved education of the non-specialist, patients and their relatives could improve recognition and outcomes in this high-risk population – including timeline access to palliative care and acute admission burden.

## Introduction

Interstitial lung diseases (ILD) represent a heterogenous group of disorders affecting the lung parenchyma, causing a spectrum of disease from inflammation to fibrosis.(1) The UK has the highest number of patients with ILD in Europe with an approximate incidence of 4.6 – 8.65 per 100,000 people per year.(2) The trajectory of disease is variable, but often results in hospitalisation secondary to respiratory decompensation. Hospital admission in ILD is associated with significant morbidity and mortality.(3)(4) A sub-set of these admissions are secondary to an acute exacerbation (AEILD). An AEILD is defined clinically by a less than 30-day deterioration in symptoms not secondary to thromboembolic, cardiac or pneumothorax events. Radiologically they are defined via high resolution computerised tomography (HRCT) demonstrating evidence of bilateral ground-glass changes superimposed on a chronic ILD.(5) Both clinical and radiological features must be present to diagnose a definite AEILD event. If only clinical criterion is met, a diagnosis of suspected AEILD can be made.(6) While AEILD was initially defined amongst a cohort of patients with idiopathic pulmonary fibrosis (IPF), it has been extrapolated more recently to encompass all ILDs. Retrospective data has estimated annual incidence of AEILD at 9%, with AEILD events accounting for an estimated 41% of ILD-related admissions.(7)(8)

It has been increasingly recognised that ILD-related hospital admissions have a significant impact on disease trajectory, with both high inpatient and 90-day mortality rates observed.(4)(8)(9)(10) However, there is limited worldwide data reporting ILD-related admission outcomes. Management of ILD-related hospitalisations and AEILD events are currently guided by international consensus statements based on expert opinion. These statements recommend supportive interventions, although the scope of recommendations are limited due to a lack of robust evidence.(6)(11) At present, only one negative randomised control trial in the treatment of AEILD has been published(12) and there is a major need to increase research capacity in AEILD.(13) There is also recognised heterogeneity amongst the international community when it comes to diagnosis and management of AEILD events – including use of CT imaging. Europe has the lowest rates of CT use in AEILD diagnosis (67% vs. 91% in Asia) but no UK-specific data is currently available.(14)

We aimed to understand the role of ILD-related hospitalisation in ILD all-cause mortality using real-world data from the North West of England. Secondary aims included understanding the risk factors associated with 90-day all-cause mortality in ILD-related hospital admissions and AEILD, alongside diagnostic and management patterns in AEILD amongst a real-world dataset. By increasing our understanding from real-world data, we can hope to identify research and service design priorities.

## Methods

### Study Setting and Population

We performed a multicentre retrospective observational cohort study of ILD-related admissions in adults ≥18 years old using International Classification of Diseases Version 10 (ICD-10) coding across a 3-year period (1^st^ January 2017 to 31^st^ December 2019) from seven NHS trusts, encompassing 11 NHS secondary and tertiary hospitals, in the North West of England, covering an estimated population of 7.3million people.(15) ICD-10 codes of B22.1, D86.0, D86.2, J67.0-67.9, J70.2-70.4, J84.1, J84.8 and J84.9 within the primary diagnosis of admission event triggered inclusion in the study (supplementary table 1). Admissions for attendance at day-case procedures, for provision of medications or lack of available medical records were excluded. Any prior documented dissent for use of data in research also resulted in exclusion (supplementary figure 1).

Medical records were retrospectively reviewed, and hand searched between June 2024 and April 2025. The primary outcome was number of days from start of ILD-related admission to death.

Ethical approval for this study was granted by the Health Research Authority (HRA) for England and Wales (HRA reference: 23/HRA/4562). Given the retrospective nature of the study and anticipated high mortality rate, individual patient consent was not required. Only patients with previously recorded consent for the use of their data in research were included. Identifiable data was accessed and processed solely by site-specific clinicians who were part of the patients’ direct care teams. All data was anonymised at the point of site-level collection, in accordance with the General Data Protection Regulation (GDPR).(16)

### Patient and Public Involvement

Rationale for developing this research question initially stemmed from the published top ten priorities for progressive pulmonary fibrosis, of which its stakeholders included patients living with PPF and their relatives and carers. Amongst the top 15 questions, two were related to acute exacerbations: a) “How can acute deteriorations of PPF be predicted in patients with PPF?” and b) “what is the best management of acute deterioration in PPF?”.(17)

Following the development of the study protocol, the concept and planned design was presented to a group of expert patients, the Research Champions through Action for Pulmonary Fibrosis, prior to IRAS submission. They confirmed acceptability of the proposed design and assisted with planning of a dissemination strategy, including radio and newsletter appearances following publication.

### Data Collection and Variables

Data collected was sourced from coding and hand searching of medical records. Where available, data collection on demographics and pre-admission diagnoses included: age, gender, postcode, ethnicity, smoking history, co-morbidities, ILD diagnosis, pre-admission antifibrotic use, pre-admission oxygen use, pre-admission lung function tests (including forced vital capacity (FVC)) and gas transfer factor (TLCO)). Comorbidities were converted to a Charlson Comorbidity Index (CCI) value.(18) Full postcodes were converted to deprivation deciles (DD), as per the 2019 UK Government English Indices of Deprivation data.(19) A DD of 1 represents the most deprived and 10 the least deprived.

Mortality-relevant data was obtained from coding and hand searching of inpatient medical records. This included: AEILD status, at-admission investigations (white cell count and differential, C-reactive protein (CRP), chest x-ray findings), data on computerised tomography (CT) of the chest, echocardiogram and bronchoscopy use, and provision of antibiotics or steroids throughout admission. Data on ventilatory support given, specialist respiratory and palliative reviews were also obtained from the notes.

Patients were allocated to AEILD or ‘other’ ILD-related admission cohorts. AEILD status was determined by hand searching of medical records and investigations relating to the admission event. For the purposes of this study, we defined AEILD clinically only by <30-day clinical deterioration, not secondary to cardiac failure, thromboembolic disease (TED) or pneumothorax event. Hand-searching of the clinical narrative, biochemical investigations (negative pro-B natriuretic peptide (pro-BNP) and negative D-dimer, where available), radiological investigations (chest x-ray and CT pulmonary angiogram (CTPA), where available) and medical treatments determined this status. If a patient met this criterion, their admission was labelled ‘AEILD’ and if not, was labelled as ‘other’ ILD-related admission. Other ILD-related admission reasons included disease progression, pulmonary embolism, cardiac failure secondary to pulmonary hypertension and pneumothorax. A respiratory review was considered a review from any respiratory-specialist healthcare professional. A palliative review was considered a review from any palliative-specialist healthcare professional.

The primary outcome was established by determining the number of days from admission start date to date of death, recorded as number of days.

### Statistical Analysis

To power the study to achieve statistical significance, we used a predicted overall all-cause mortality of 60% in the follow-up period, a mortality-based model of ten predictors and an R-squared of 0.1 determined the inclusion of 850 admission events. We chose a multicentre, 3-year inclusion period to achieve this threshold.

All statistical analysis was undertaken using SPSS v31.0 and Python v3.9.7., with the ‘statsmodel’ package for Cox proportional hazards modelling. Continuous observational data was analysed by mean (standard deviation). Categorical observational data was reported as frequencies (percentage). Admissions were grouped into AEILD or other primary admission based on clinical criterion defined above. For sub-analysis of the AEILD cohort, patients were grouped by presence of CT-confirmed AEILD (definite AEILD) and absence of CT-confirmed AEILD (suspected AEILD). Continuous data was assessed for normality using the Shapiro-Wilks test and then compared using the unpaired t-test or Mann-Whitney-U test dependent on presence of normal distribution. Categorical data was compared with the chi squared test or Fisher’s exact test if the expected count was less than five. Post-hoc analysis of chi-squared with multiple comparisons was undertaken using standardised residuals, converted to P values and compared to the Bonferroni correction for statistical significance.

For time-to-event outcomes, Kaplan Meier survival analysis with log-rank comparison for 2 or more curves was undertaken. Mean and median survival were calculated. To quantify risk for all-cause mortality, hazard ratios with corresponding 95% confidence intervals were estimated using a multivariate Cox proportional hazards model using the ENTER method. Variables were selected and entered into the model simultaneously. Variable selection was informed by both clinical relevance and prior evidence, rather than data-driven stepwise methods, to minimise model overfitting. Demographic characteristics (e.g. age, gender, ethnicity) were included to assess the impact of non-modifiable risk factors, while clinical and inflammatory variables were chosen for their known or hypothesised relevance to mortality risk in ILD populations.(20)(21)(22) For AEILD cohort sub-analysis, CT confirmation status and treatment strategies were also included in the model. The proportional hazards assumption was checked using Schoenfeld residuals. Results yielding p <0.05 were considered statistically significant.

Complete case analysis was undertaken. Variables with high proportions of missing data (>15%), once data collection had been undertaken, were removed. FVC was originally planned to be included in the model but removed due to a high proportion of missing data (390/938 or 38.9%).

## Results

3451 admissions were identified. 2449/3451 (70.9%) of admissions were excluded. Main reason for exclusion was day case procedure (1984/2449, 81.1%, supplementary figure 1). The remaining 1002 admission events underwent hand-searching of medical records and coding data. 64/1002 (6.4%) had insufficient data available to determine AEILD vs. other ILD-related admission status, leaving 938 admissions for inclusion in analysis. Median follow-up amongst censored patients (those alive at the end of the study period) was 2125 days (range 1563 – 2968 days).

### Descriptive Statistics

In total, 511/938 (54.5%) of admission events met AEILD criteria as per the study definition and 427/938 (45.5%) were considered other ILD-related admissions.

Baseline and admission characteristics, including rates of CT confirmation, respiratory and palliative specialist review and treatments, for the overall dataset and AEILD sub-group analysis are summarised in tables 1 and 2 respectively.

**Table 1:** Summary of descriptive patient characteristics for overall, acute exacerbation of interstitial lung disease (AEILD) and other ILD-related admission cohorts. P-value refers to comparison between AEILD and other cohorts using independent T-test or Mann-Whitney-U test for continuous variables, and chi-squared or Fisher’s exact test for categorical variables. Post-hoc analysis of chi-squared was undertaken using standardised residuals, converted to P values and compared to the Bonferroni correction for statistical significance. *Abbreviations: AE-ILD – acute exacerbation of ILD; CCI – Charlson Comorbidity Index; ILD – Interstitial Lung Disease; IPF – Idiopathic pulmonary fibrosis; NSIP – non-specific interstitial pneumonitis; CTD-ILD – connective tissue disease interstitial lung disease; HP – hypersensitivity pneumonitis; PFD – primary fibrosis as diagnostic label; SD – standard deviation; CT – computerised tomography*

|  | Overall<br>n = 938 | AEILD Cohort<br>n = 511 | Other Cohort<br>n = 427 | P value | Adjusted P value<br>(Bonferroni) |
| --- | --- | --- | --- | --- | --- |
| **Baseline Characteristics** |  |  |  |  |  |
| **Age**<br>(Mean, SD) | 74.18 (12.48) | 73.87 (11.63) | 74.55 (13.42) | p = 0.058 |  |
| **Gender – Male**<br>(Number, %) | 521 (55.5) | 288 (56.4) | 233 (54.6) | p = 0.582 |  |
| **Ethnicity**<br>(Number, %) |  |  |  | p = 0.067 |  |
| Not stated | 61 (6.5) | 25 (4.9) | 36 (8.4) | **p = 0.029\*** | p = 0.115 |
| White | 802 (85.5) | 444 (86.9) | 358 (83.8) | p = 0.187 |  |
| Asian | 68 (7.2) | 40 (7.8) | 28 (6.6) | p = 0.455 |  |
| Black | 7 (0.7) | 2 (0.4) | 5 (1.2) | p = 0.167 |  |
| **CCI**<br>(Mean, SD) | 4.46 (1.98) | 4.38 (1.86) | 4.54 (2.11) | p = 0.165 |  |
| **Deprivation Decile**<br>(Mean, SD) | 4.62 (2.89) | 4.62 (2.92) | 4.63 (2.85) | p = 0.980 |  |
| **Ever-smoker**<br>(Number, %) | 488 (52.0) | 262 (51.3) | 226 (52.9) | p = 0.786 |  |
| **ILD Subtype**<br>(Number, %) |  |  |  | **p <0.0001\*** |  |
| Not stated | 91 (9.7) | 22 (4.3) | 69 (16.2) | **p <0.0001\*** | **p <0.0001\*** |
| IPF | 295 (31.4) | 178 (34.8) | 117 (27.4) | p = 0.015 | p = 0.161 |
| NSIP | 84 (9.0) | 52 (10.2) | 32 (7.5) | p = 0.152 |  |
| CTD-ILD | 46 (4.9) | 28 (5.5) | 18 (4.2) | p = 0.372 |  |
| HP | 77 (8.2) | 44 (8.6) | 33 (7.7) | p = 0.624 |  |
| Drug-related | 22 (2.3) | 9 (1.8) | 13 (3.0) | p = 0.196 |  |
| Industry-related | 19 (2.0) | 10 (2.0) | 9 (2.1) | p = 0.870 |  |
| Sarcoidosis | 25 (2.7) | 11 (2.2) | 14 (3.3) | p = 0.286 |  |
| PFD | 221 (23.6) | 126 (24.7) | 95 (22.2) | p = 0.387 |  |
| Unclassifiable | 28 (3.0) | 18 (3.5) | 10 (2.3) | p = 0.290 |  |
| Other | 30 (3.2) | 13 (2.5) | 17 (4.0) | p = 0.213 |  |
| **On antifibrotics** | 118 (12.6) | 73 (14.3) | 45 (10.5) | **p = 0.001\*** |  |
| *(Number, %)* |  |  |  |  |  |
| **Oxygen used pre-admission**<br>(Number, %) |  |  |  | **p = 0.013\*** |  |
| Long-term | 256 (27.3) | 153 (29.9) | 103 (24.1) | p = 0.046 | p = 0.139 |
| Ambulatory | 78 (8.3) | 50 (9.8) | 28 (6.6) | p = 0.075 | p = 0.224 |
| None | 604 (64.4) | 308 (60.3) | 296 (69.3) | p = 0.004 | **p = 0.012\*** |
| **FVC (L)**<br>(Mean, SD) | 2.11 (0.83) | 2.16 (0.86) | 2.03 (0.79) | p = 0.102 |  |
| **TCLO (mmHg)**<br>(Mean, SD) | 3.88 (2.19) | 3.37 (1.61) | 4.64 (2.67) | **p <0.0001\*** |  |
| **Admission Characteristics** |  |  |  |  |  |
| **Any CT of thorax performed**<br>Number (%) | 326 (34.8) | 192 (37.6) | 134 (31.4) | p = 0.05 |  |
| **Respiratory Specialist Review**<br>Number (%) | 508 (54.2) | 350 (68.5) | 158 (37.0) | **p = <0.0001\*** |  |
| **Palliative Specialist Review**<br>Number (%) | 130 (13.9) | 92 (18.0) | 38 (8.9) | **p = 0.0003\*** |  |
| **Steroids**<br>Number (%) |  |  |  | **p <0.0001\*** |  |
| **Methylprednisolone** |  |  |  |  |  |
| Methylprednisolone 500mg | 14 (1.5) | 14 (2.7) | 0 (0.0) | **p = 0.0006\*** | **p = 0.002\*** |
| Methylprednisolone 1000mg | 23 (2.5) | 20 (3.9) | 3 (0.7) | **p = 0.002\*** | **p = 0.005\*** |
| None | 862 (91.9) | 460 (90.0) | 402 (94.1) | **p <0.0001\*** | **p <0.0001\*** |
| **Oral Steroids** |  |  |  | **p <0.0001\*** |  |
| Prednisolone >50mg | 19 (2.1) | 14 (2.8) | 5 (1.2) | p = 0.087 | p = 0.523 |
| Prednisolone 30-40mg | 390 (42.8) | 282 (55.5) | 108 (25.3) | **p <0.0001\*** | **p <0.0001\*** |
| Prednisolone 15-20mg | 51 (5.6) | 33 (6.5) | 18 (4.2) | p = 0.126 |  |
| Prednisolone <10mg | 12 (1.3) | 5 (1.0) | 7 (1.6) | p = 0.375 |  |
| None | 422 (46.3) | 153 (30.1) | 268 (62.8) | **p <0.0001\*** | **p <0.0001\*** |
| **Initial Antibiotics**<br>Number (%) |  |  |  | **p <0.0001\*** |  |
| Intravenous | 325 (34.6) | 231 (45.2) | 109 (25.5) | **p <0.0001\*** | **p <0.0001\*** |
| Oral | 288 (30.7) | 179 (35.0) | 94 (22.0) | **p = 0.002\*** | **p = 0.007\*** |
| None | 297 (31.7) | 91 (17.8) | 206 (48.2) | **p <0.0001\*** | **p <0.0001\*** |
| **HFNO**<br>Number (%) | 54 (5.8) | 43 (8.4) | 11 (2.6) | **p = 0.0001\*** |  |
| **CPAP or NIV**<br>Number (%) | 16 (1.7) | 15 (2.9) | 1 (0.2) | **p = 0.002\*** |  |
| **Invasive Ventilation**<br>Number (%) | 2 (0.2) | 0 (0.0) | 2 (0.5) | p = 0.607 |  |

**Table 2:**
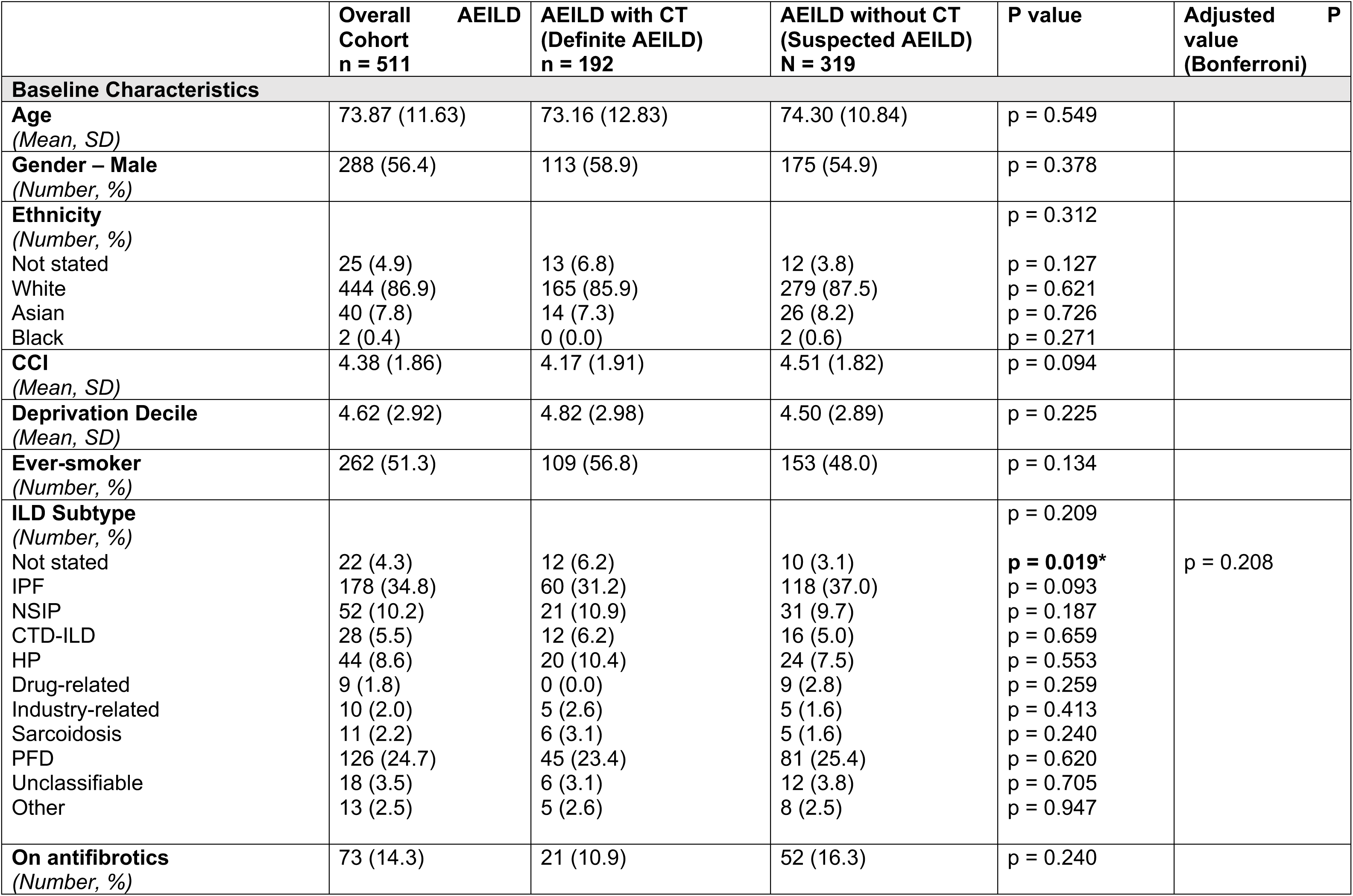
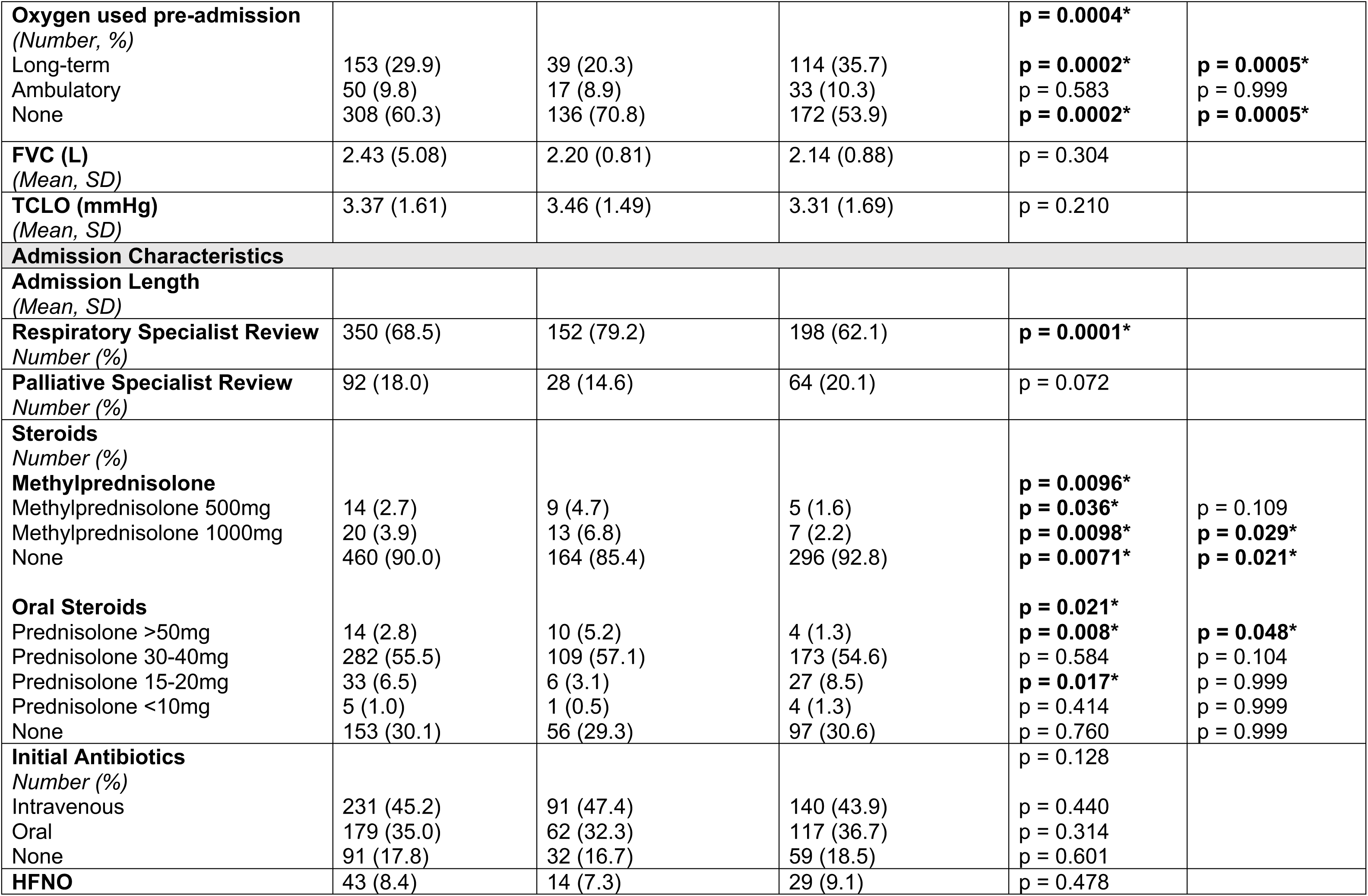
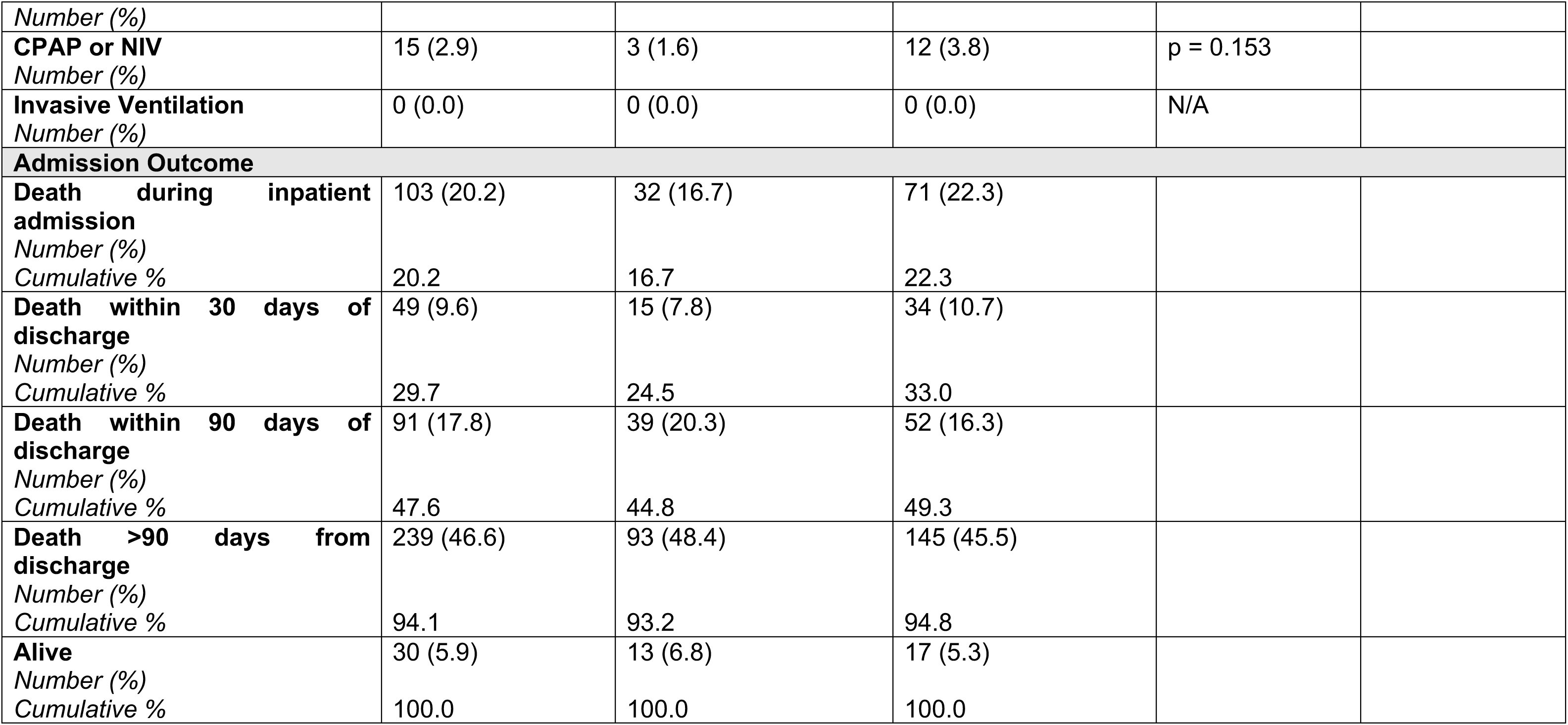
Summary of descriptive patient characteristics for all acute exacerbations (AEILD), AEILD with CT confirmation (definite AEILD) and AEILD without CT confirmation (suspected AEILD). P-value refers to comparison between AEILD and other cohorts using independent T-test or Mann-Whitney-U test for continuous variables, and chi-squared or Fisher’s exact test for categorical variables. Post-hoc analysis of chi-squared was undertaken using standardised residuals, converted to P values and compared to the Bonferroni correction for statistical significance. *Abbreviations: AE-ILD – acute exacerbation of ILD; CCI – Charlson Comorbidity Index; ILD – Interstitial Lung Disease; IPF – Idiopathic pulmonary fibrosis; NSIP – non-specific interstitial pneumonitis; CTD-ILD – connective tissue disease interstitial lung disease; HP – hypersensitivity pneumonitis; PFD – primary fibrosis as diagnostic label; SD – standard deviation; CT – computerised tomography*

### Primary Outcome – Mortality Rate and Time-to-Event Analysis

The overall 90-day mortality for this cohort was 40.2% (377/938). For the AEILD cohort, 90-day all-cause mortality was 47.6% (243/511). For the other ILD-related admission cohort, 90-day all-cause mortality was 31.4% (134/427).

Median and mean survival of AEILD cohort was 107 days (95% CI 87.0 – 141.0 days) and 408.6 days (95% CI 355.9 – 465.3 days) respectively. Median and mean survival of the other cohort was 241.0 days (95% CI 208.0 – 308.0 days) and 621.1 days (95% CI 549.5 – 700.3 days) respectively. The comparative difference survival between the two groups was statistically significant (p <0.0001; figure 1).

**Figure 1:**
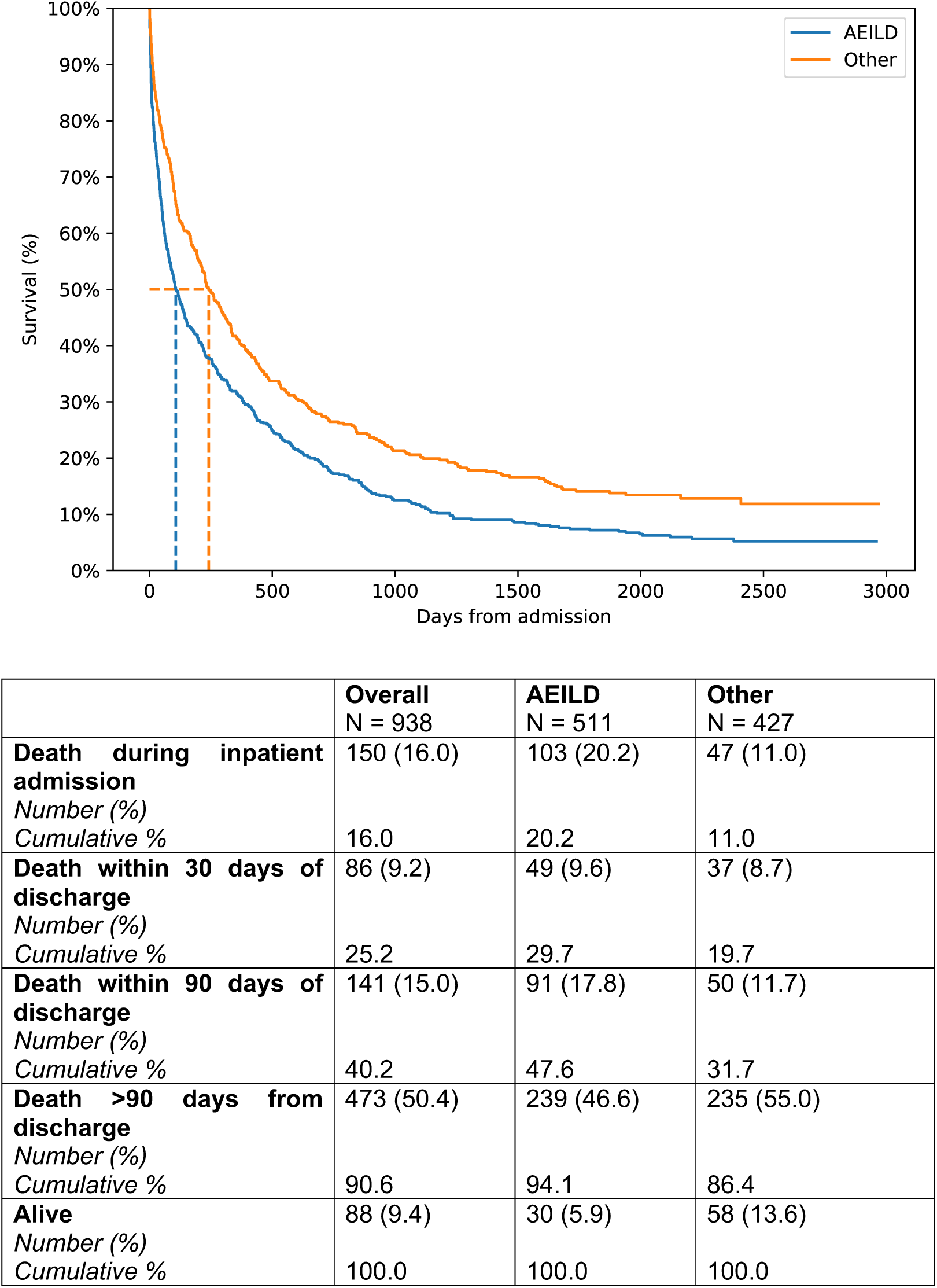
Kaplan-Meier analysis of interstitial lung disease-related admissions, comparing entire AEILD cohort (encompassing definite AEILD, with CT confirmation, and suspected AEILD, with clinical definition only met). Median and mean survival of AEILD cohort was 107 days (95% CI 87.0 – 141.0 days) and 408.6 days (95% CI 355.9 – 465.3 days) respectively. Median and mean survival of the other cohort was 241.0 days (95% CI 208.0 – 308.0 days) and 621.1days (95% CI 549.5 – 700.3 days) respectively. There was a statistically significant difference in survival between the cohorts with log-rank comparison (p <0.0001). *Abbreviations: AEILD – acute exacerbations of interstitial lung disease; other – other interstitial lung disease-related admission reason*.

### Secondary Outcomes

Ten variables were included in a multivariate model for all-cause mortality across the ILD-related hospital admissions (table 3).

**Table 3:**
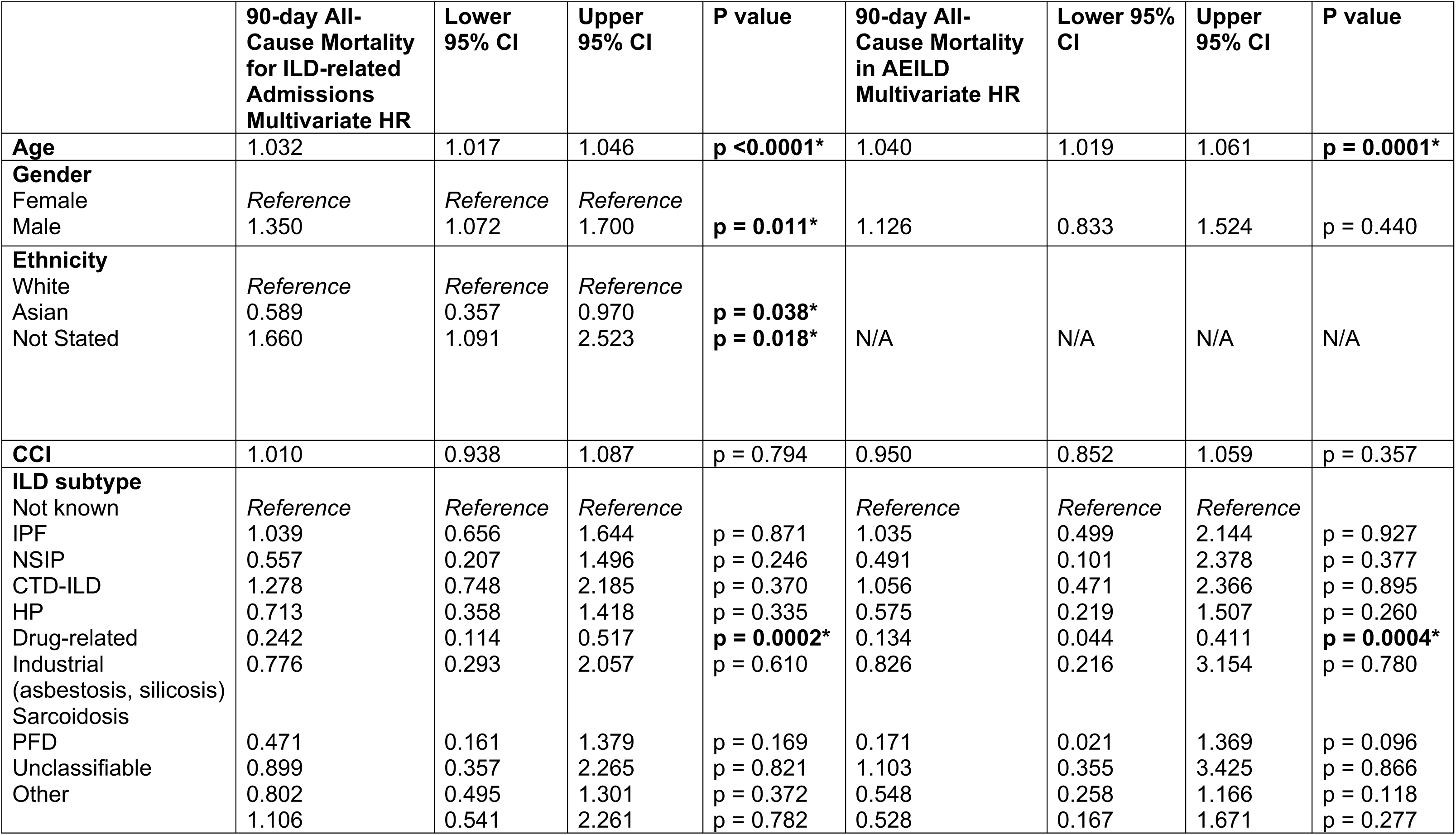
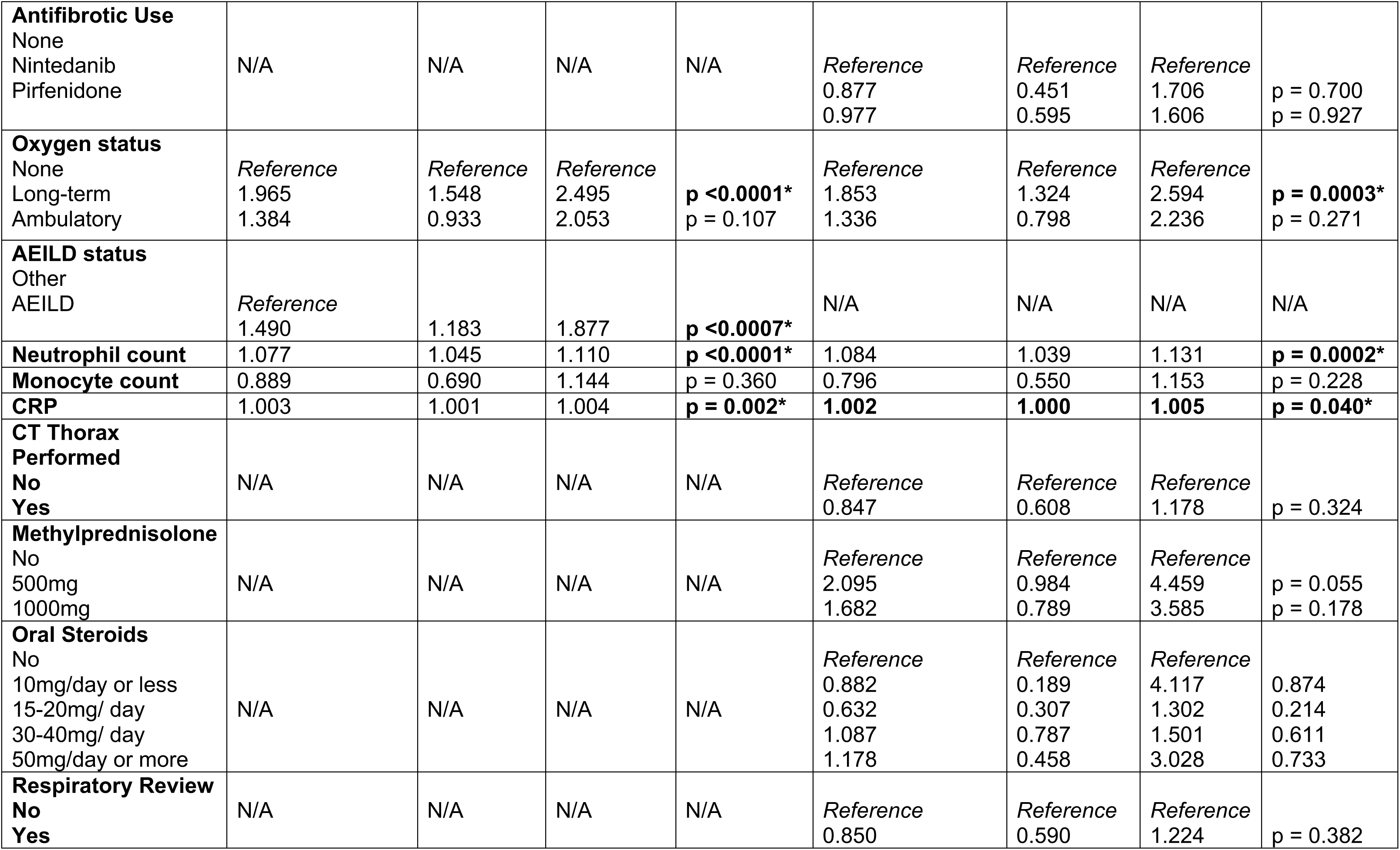
Summary table of multivariate Cox regression analysis on 90-day all-cause mortality for all ILD-related admissions and AEILD sub-group analysis. Statistically significant results are marked in bold and with an asterisk. *Abbreviations: HR – hazard ratio; CCI –Charlson comorbidity index; ILD – interstitial lung disease; AEILD – acute exacerbation of interstitial lung disease; IPF – idiopathic pulmonary fibrosis; NSIP – non-specific interstitial pneumonitis; CTD-ILD – connective tissue disease interstitial lung disease; HP – hypersensitivity pneumonitis; PFD – primary fibrosis as diagnostic label; CRP – C reactive protein*

|  | 90-day All-Cause Mortality for ILD-related Admissions Multivariate HR | Lower 95% CI | Upper 95% CI | P value | 90-day All-Cause Mortality in AEILD Multivariate HR | Lower 95% CI | Upper 95% CI | P value |
| --- | --- | --- | --- | --- | --- | --- | --- | --- |
| **Age** | 1.032 | 1.017 | 1.046 | **p <0.0001\*** | 1.040 | 1.019 | 1.061 | **p = 0.0001\*** |
| **Gender** |  |  |  |  |  |  |  |  |
| Female | *Reference* | *Reference* | *Reference* |  |  |  |  |  |
| Male | 1.350 | 1.072 | 1.700 | **p = 0.011\*** | 1.126 | 0.833 | 1.524 | p = 0.440 |
| **Ethnicity** |  |  |  |  |  |  |  |  |
| White | *Reference* | *Reference* | *Reference* |  |  |  |  |  |
| Asian | 0.589 | 0.357 | 0.970 | **p = 0.038\*** |  |  |  |  |
| Not Stated | 1.660 | 1.091 | 2.523 | **p = 0.018\*** | N/A | N/A | N/A | N/A |
| **CCI** | 1.010 | 0.938 | 1.087 | p = 0.794 | 0.950 | 0.852 | 1.059 | p = 0.357 |
| **ILD subtype** |  |  |  |  |  |  |  |  |
| Not known | *Reference* | *Reference* | *Reference* |  | *Reference* | *Reference* | *Reference* |  |
| IPF | 1.039 | 0.656 | 1.644 | p = 0.871 | 1.035 | 0.499 | 2.144 | p = 0.927 |
| NSIP | 0.557 | 0.207 | 1.496 | p = 0.246 | 0.491 | 0.101 | 2.378 | p = 0.377 |
| CTD-ILD | 1.278 | 0.748 | 2.185 | p = 0.370 | 1.056 | 0.471 | 2.366 | p = 0.895 |
| HP | 0.713 | 0.358 | 1.418 | p = 0.335 | 0.575 | 0.219 | 1.507 | p = 0.260 |
| Drug-related | 0.242 | 0.114 | 0.517 | **p = 0.0002\*** | 0.134 | 0.044 | 0.411 | **p = 0.0004\*** |
| Industrial | 0.776 | 0.293 | 2.057 | p = 0.610 | 0.826 | 0.216 | 3.154 | p = 0.780 |
| (asbestosis, silicosis) |  |  |  |  |  |  |  |  |
| Sarcoidosis |  |  |  |  |  |  |  |  |
| PFD | 0.471 | 0.161 | 1.379 | p = 0.169 | 0.171 | 0.021 | 1.369 | p = 0.096 |
| Unclassifiable | 0.899 | 0.357 | 2.265 | p = 0.821 | 1.103 | 0.355 | 3.425 | p = 0.866 |
| Other | 0.802 | 0.495 | 1.301 | p = 0.372 | 0.548 | 0.258 | 1.166 | p = 0.118 |
|  | 1.106 | 0.541 | 2.261 | p = 0.782 | 0.528 | 0.167 | 1.671 | p = 0.277 |
| **Antifibrotic Use**<br>None<br>Nintedanib<br>Pirfenidone | N/A | N/A | N/A | N/A | Reference<br>0.877<br>0.977 | Reference<br>0.451<br>0.595 | Reference<br>1.706<br>1.606 | p = 0.700<br>p = 0.927 |
| **Oxygen status**<br>None<br>Long-term<br>Ambulatory | Reference<br>1.965<br>1.384 | Reference<br>1.548<br>0.933 | Reference<br>2.495<br>2.053 | p < 0.0001*<br>p = 0.107 | Reference<br>1.853<br>1.336 | Reference<br>1.324<br>0.798 | Reference<br>2.594<br>2.236 | p = 0.0003*<br>p = 0.271 |
| **AEILD status**<br>Other<br>AEILD | Reference<br>1.490 | 1.183 | 1.877 | p < 0.0007* | N/A | N/A | N/A | N/A |
| **Neutrophil count** | 1.077 | 1.045 | 1.110 | p < 0.0001* | 1.084 | 1.039 | 1.131 | p = 0.0002* |
| **Monocyte count** | 0.889 | 0.690 | 1.144 | p = 0.360 | 0.796 | 0.550 | 1.153 | p = 0.228 |
| **CRP** | 1.003 | 1.001 | 1.004 | p = 0.002* | 1.002 | 1.000 | 1.005 | p = 0.040* |
| **CT Thorax Performed**<br>No<br>Yes | N/A | N/A | N/A | N/A | Reference<br>0.847 | Reference<br>0.608 | Reference<br>1.178 | p = 0.324 |
| **Methylprednisolone**<br>No<br>500mg<br>1000mg | N/A | N/A | N/A | N/A | Reference<br>2.095<br>1.682 | Reference<br>0.984<br>0.789 | Reference<br>4.459<br>3.585 | p = 0.055<br>p = 0.178 |
| **Oral Steroids**<br>No<br>10mg/day or less<br>15-20mg/ day<br>30-40mg/ day<br>50mg/day or more | N/A | N/A | N/A | N/A | Reference<br>0.882<br>0.632<br>1.087<br>1.178 | Reference<br>0.189<br>0.307<br>0.787<br>0.458 | Reference<br>4.117<br>1.302<br>1.501<br>3.028 | 0.874<br>0.214<br>0.611<br>0.733 |
| **Respiratory Review**<br>No<br>Yes | N/A | N/A | N/A | N/A | Reference<br>0.850 | Reference<br>0.590 | Reference<br>1.224 | p = 0.382 |

Sub-analysis of the AEILD cohort was undertaken through assigning patients to definite or suspected AEILD by presence of CT imaging confirmation. 37.6% (192/511) of AEILD admissions had CT thorax imaging to confirm diagnosis. Mean and median survival of AEILD with CT confirmation was 476.8 days (95% CI 369.0 – 566.1) and 144.0 days (95% CI 87.0 – 237.0). Mean and median survival of AEILD without CT confirmation was 359.3 days (95% 289.1 days – 425.2 days) and 100.0 days (95% CI 71.0 – 132.0), with a statistically significant difference between the two cohorts (p = 0.027; figure 2). Multivariate modelling of 90-day all-cause mortality for the AEILD sub-group was also undertaken with 13 variables (table 3).

**Figure 2:**
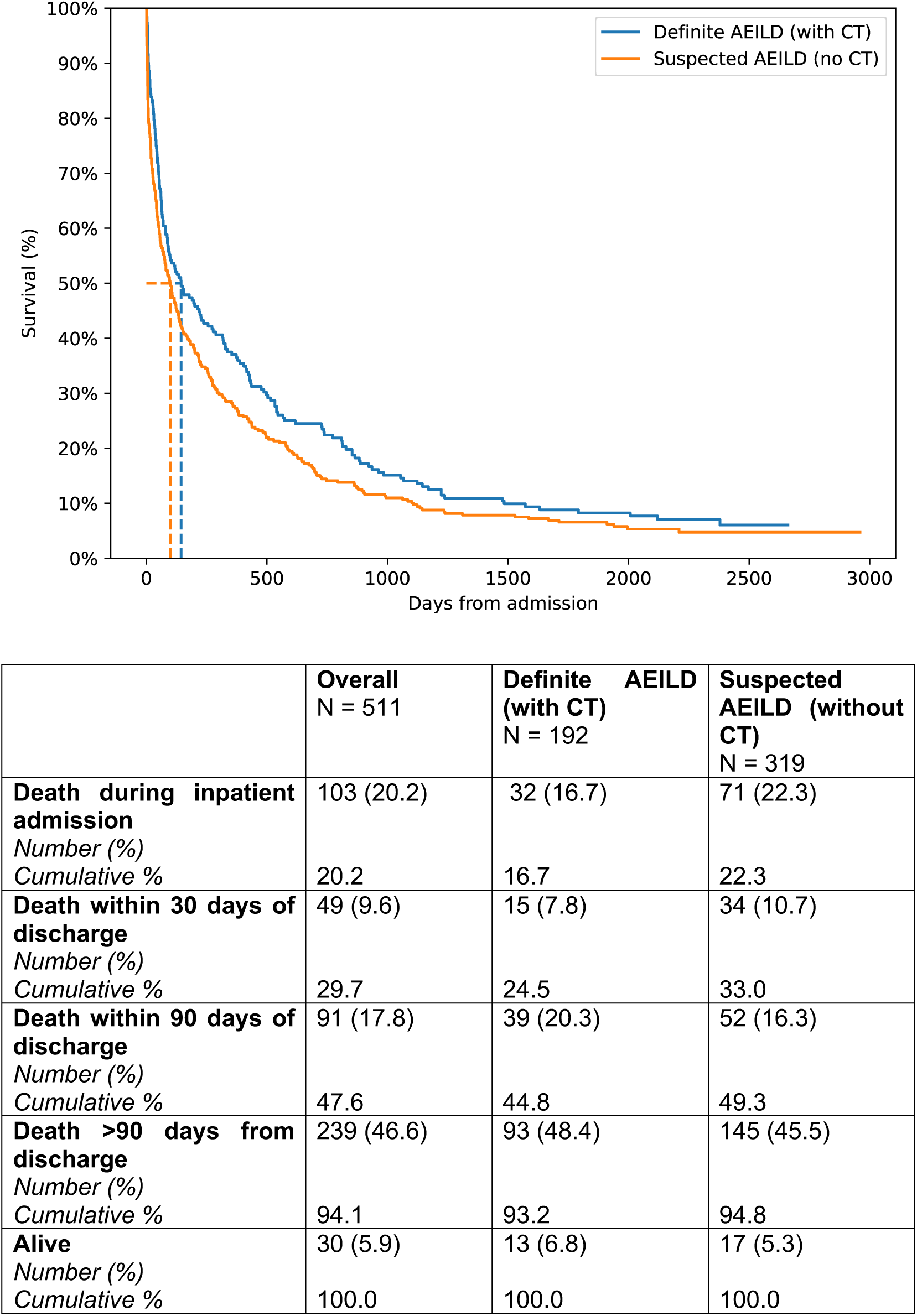
Kaplan-Meier analysis of AEILD cohort, comparing definite AEILD (with CT confirmation) versus suspected AEILD (without CT confirmation) using log-rank analysis. Mean and median survival of AEILD with CT was 476.8 days (95% CI 369.0 – 566.1) and 144.0 days (95% CI 87.0 – 237.0). Mean and median survival of AEILD without CT was 359.3 days (95% 289.1 days – 425.2 days) and 100.0 days (95% CI 71.0 – 132.0), with a statistically significant difference between the two cohorts (p = 0.027). *Abbreviations: AEILD – acute exacerbations of interstitial lung disease; CT – computerised tomography*.

## Discussion

### Summary of Findings

To the best of our knowledge, this is the largest real-world UK dataset analysing ILD-related admission outcomes. Our data demonstrates significant 90-day all-cause mortality across all ILD-related admissions (40.2%) – but this mortality is significantly greater in an AEILD admission compared to other ILD-related admission reasons (median survival 107 days vs. 241 days; p <0.0001). AEILD remained a risk factor for mortality in a multivariate model of 90-day mortality (HR 1.490, 95% CI 1.183–1.877; p = 0.0007), alongside pre-admission long-term oxygen use (HR 1.965, 95% CI 1.548–2.495; p <0.0001), age (HR 1.032, 95% CI 1.017-1.046; p <0.0001), male gender (HR 1.350, 95% CI 1.072–1.700; p = 0.011) and neutrophilia (HR 1.077, 95% CI 1.045–1.110; p <0.0001). Ethnicity demonstrated a variable association, with Asian ethnicity (compared to a reference of White ethnicity) demonstrating protection against mortality (HR 0.589, 95% CI 0.357–0.970; p = 0.038), whereas not stating an ethnicity increased mortality (HR 1.660, 95% CI 1.091 2.523; p = 0.018). Drug-related ILD was also associated with protection against mortality (HR 0.242, 95% CI 0.114–0.517; p = 0.0002).

Given our AEILD cohort encompassed both definite AEILD (with CT confirmation) and suspected AEILD (without CT confirmation), sub-analysis was undertaken to understand the impact of CT confirmation on outcomes. Our data demonstrates there was a statistically significant difference in mortality between those with and without CT confirmation (median survival 144 days vs. 100 days; p = 0.027). Interestingly, it was our group without CT confirmation who experienced higher mortality. When taken into a multivariate model for all AEILD admissions, CT confirmation was not significantly associated with reduced risk of mortality (HR 0.847, 95% CI 0.608 – 1.178; p = 0.324). Age, pre-admission oxygen and neutrophilia were again associated with increased risk of mortality, and drug-related ILD reduced risk. Methylprednisolone and high-dose steroid use (>50mg/day) also demonstrated hazard ratios suggesting increased risk of mortality within the model – but did not meet statistically significant thresholds.

These findings underscore the high mortality burden associated with ILD-related admissions and highlight diagnostic and management heterogeneity that may contribute to outcome variation.

### Study Strengths and Limitations

The main strength of this study is its multicentre design, encompassing several geographical regions in the North-West of England – enhancing its power. By also including a mix of secondary and tertiary hospitals, these combined strengths increase the generalisability of the study – but key limitations remain.

Our retrospective data is between 2017 and 2019. While this timeframe was chosen to remove the confounding variable of the COVID-19 pandemic, it is important to recognise the change in the treatment landscape in the period since this data collection, such as wider antifibrotic use. As such, the study may not reflect current patient outcomes.

The retrospective design also introduces the potential for recall and misclassification bias. This was particularly important for determining AEILD based on clinical, biochemical and radiological information available. While hand-searching notes and investigations aimed to mitigate misclassification bias, inter-rate reliability was not assessed, and full CT reports were not recorded – which may affect consistency in classification. Missing data is also a further challenge of retrospective data, and we observed high rates of this when recording lung function testing (FVC and TLCO specifically). This high rate of missing data meant the need to remove FVC from our multivariate model. Additionally, hospital-only based data collection risks hospitalisation bias. Different results may be observed if respiratory decompensation events relating to ILD that don’t result in hospital admission are also accounted for.

Despite covering a large geographic area in the North West of England, this study is region-specific and may not generalise to different health systems or ILD pathways observed outside of it. Further studies on a national scale, ideally with prospective data collection, are required to validate the results observed here.

### The Role of AEILD in ILD Admission Mortality

AEILD events are not infrequent in the hospitalised ILD population. A prior retrospective study identifying AEILD events with clinical criterion only reported an AEILD rate of 52%(23) -similar to the rate we observed. In studies where radiological and clinical criterion were used, fulfilling “definite AEILD” criteria set out by Collard el al,(6) the rate of AEILD in hospitalised patients was 20.8% -41%.(8)(24)

Not only are AEILDs frequent events, but they are associated with high mortality. Prior literature has reported mortality of up to 100% in the intensive care setting.(25) Our findings reinforce this high mortality amongst a UK population: AEILD admissions were associated with significantly higher 90-day mortality compared with other ILD-related admissions. Notably, median survival following AEILD admission was only 107 days and remained significantly different to other ILD-related admission reasons – even after multivariate adjustment. This suggests an AEILD event itself represents a distinct and high-risk clinical picture. However, there are a proportion of patients who survive beyond the 90-day mark for considerable periods (figure 1), raising the possibility of different phenotypes, causes and severities within the umbrella term AEILD. The possibility of differing AEILD severities was addressed in a post-hoc analysis for exacerbators within the INPULSIS trial, with those meeting criteria for a serious adverse event demonstrating worse outcomes.(26)

The hypothesis of differing AEILD phenotypes, causes and severities is also supported by the observed differences in AEILD outcomes between differing ILD underlying disease types,(27) which is further suggested in our multivariate model – although not to a statistically significant degree. The relatively small numbers within each ILD sub-group may have impacted this dataset’s ability to reach statistically significant thresholds. It is likely also limited by the choice to group by disease, rather than radiological pattern. The 2025 ERS/ATS guidelines have moved initial diagnostic labelling to a two-step process, with the first being based on morphological patterns by pathology and imaging.(28) This distinction appears important in AEILD, too, with usual interstitial pneumonia (UIP) and nonspecific interstitial pneumonitis (NSIP) radiological patterns shown to have different survival outcomes.(29)(30) Future studies of this work should therefore look to categorising by radiological pattern, rather than disease, to understand the impact on mortality outcomes more clearly.

The mechanisms driving poor outcomes in AEILD are poorly understood,(5) and treatment options remain limited.(11)(13) The current international consensus guidance recommends supportive care but lacks specific pharmacological and non-pharmacological strategies.(6) Our results support the argument that AEILD represents not only an acute crisis but also a turning point in the disease course. The elevated mortality risk underscores the need for early identification, prompt multidisciplinary input, and targeted therapeutic trials in this population alongside clear, defined, palliative and end-of-life care strategies.

### Impact of CT Confirmation on AEILD Outcomes

CT thorax imaging currently remains an essential part of the diagnostic criteria for confirming an AEILD.(6) However, there is heterogeneity amongst its use internationally(14) – and patterns of its use have never been studied in a specific UK dataset.

In our real-world dataset, we demonstrate poor uptake of CT imaging to confirm an AEILD diagnosis. This was also associated with a significant difference in survival, whereby mortality risk increased if CT confirmation was not undertaken (also referred to as suspected AEILD) in Kaplan-Meier survival analysis. There are some potential explanations for this. First, patients with AEILD often develop profound respiratory failure(31) – meaning they may not be safe to transfer to CT imaging to confirm the diagnosis. However, this data did not demonstrate significant differences in high flow nasal oxygen (HFNO) use, non-invasive ventilation (NIV) nor specialist palliative care inpatient review. A second possibility is that CT is a marker for more intensive therapies and specialist care. The group with CT did demonstrate a higher respiratory review rate (62.1% in suspected AEILD vs. 79.2% in definite AEILD; p = 0.0001; table 2) and higher rate of methylprednisolone (1.6% in suspected AEILD vs. 4.7% in definite AEILD for 500mg; p = 0.036; 2.2% in suspected AEILD vs. 6.8% in definite AEILD for 1000mg; p = 0.0098; table 2) or high-dose (>50mg/day) prednisolone use (1.3% in suspected AEILD vs. 5.2% in definite AEILD, p = 0.008; table 2). However, in a multivariate model of 90-day AEILD all-cause mortality, neither CT, nor respiratory review, nor steroid use met statistically significant thresholds for association with or protection from mortality. In fact, high-dose steroid use had numerical hazard ratios associated with increased mortality (table 3) – but this may be confounded by the fact the most unwell patients receive this therapy.(32) Instead, increased age, pre-admission long-term oxygen therapy, and neutrophilia were significantly associated with increased all-cause mortality across all AEILD admissions.

There is limited data comparing outcomes from suspected vs. definite AEILD events. A small post-hoc analysis from the STEP-IPF trial found no significant difference in mortality between suspected and definite AE-IPF events – suggesting both convey significant post-event mortality.(33) Our findings support this, with a multivariate model highlighting the complex interaction of many variables contributing to AEILD mortality.

These findings have important service and education implications. They emphasise the need for harmonised diagnostic criteria and greater accessibility of HRCT in the acutely unwell population, to ensure consistent identification and management of AEILD events. If CT remains an essential part of the diagnostic criteria, then the low rate of CT use also highlights an essential education need for the non-respiratory physician who may see these patients in the emergency department, or on the acute medical take. They also raise the question of the possible need for an inclusive, real-world diagnostic criteria that may not necessitate CT imaging – but these findings first need confirmation in a validatory cohort.

### Palliative Care Input

Our study highlights a concerning underutilisation of specialist palliative care services in a population with exceptionally high short and long-term mortality – greater than many common cancers. Less than 1 in 5 patients experiencing an AEILD received inpatient specialist palliative care reviews despite a 90-day mortality of just under 50%. There have been several calls to improve access and use of palliative care services in ILD, with a growing understanding of the unmet needs of this population.(34)(35)(36) But a lower referral rate may reflect the clinical uncertainty ILD brings and the lack of direct palliative recommendations in the context of a respiratory decompensation or AEILD event.(6)(37) This reflects the lack of evidence in this area, and, much of what is available being of low quality to base such recommendations on.(6) There is also a well-recognised international heterogeneity in approaches to AEILDs, with 50-71% of specialist clinicians reporting they routinely offer palliative care support in the context of ILD.(14)

In the UK, the National Institute for Clinical Excellence (NICE) care quality standard 5 for idiopathic pulmonary fibrosis (IPF) states: “people with IPF and their families and carers have access to services that meet their palliative care needs”.(38) The recent British Thoracic Society (BTS) ILD registry report summarising 2024 data demonstrates the heterogeneity in meeting this care standard across UK hospitals,(39) further supported by our dataset findings. However, our study may underappreciate the palliative care provided by respiratory and general physicians. Our study recorded palliative care input as a documented inpatient review by a palliative care specialist only. More recent palliative-focused studies have shown the merits of a multidisciplinary team (MDT) model to improve patient-reported outcomes.(40) Follow-up studies should acknowledge palliative care input from the non-palliative specialist to build a full picture of the palliative care approaches in ILD admissions.

This current underutilisation of palliative care in the context of ILD-related admissions and AEILD highlight again an area of essential education for the non-specialist. Through education and improved awareness of poor prognosis in these events, alongside improved diagnostic confirmation, may help to improve patient and carer access to the vital palliative and supportive care services they need.

### The Ethnic Paradox

The impact of ethnicity on ILD outcomes has not yet been established. In a US population, ethnic disparities were observed across 4792 participants with pulmonary fibrosis, with black ethnicity associated with diagnosis at a younger age – but paradoxically lower mortality rates.(41) In our dataset, compared to White ethnicity, Asian ethnicity was associated with reduced risk of 90-day all-cause mortality. In UK epidemiology, this is referred to as the “ethnic paradox”: where some ethnic minority groups, especially of South Asian background, have higher burden of chronic disease but paradoxically lower mortality rates.(42) Several other studies mirror this finding of reduced mortality in ethnic minorities,(43) suggesting the complex interaction of social determinants of health, health-seeking behaviours, biological differences and cultural practices. Future studies should prioritise complete and consistent ethnicity data collection and be powered to explore subgroup differences meaningfully.

## Conclusion

This retrospective study demonstrates high mortality in the 90 days following ILD-related inpatient admissions, especially in the context of AEILD. It demonstrates the real-world application of the AEILD diagnostic criteria, highlighting essential need for education amongst the non-ILD specialist to improve rates of CT use and timely palliative care utilisation.

## Acknowledgements

Thank you to the Northern Care Alliance NHS Foundation Trust for providing sponsorship of the study.

Thank you to the National Institute of Health and Social Care Research for the Academic Clinical Fellowship programme which afforded LW protected time to conduct this study.

Thank you to Professor Andy Vail for providing statistical guidance.

## Disclaimers

The views expressed in the submitted article are our own and not an official position of the affiliated institutions or the Northern Care Alliance NHS Foundation Trust.

## Contributorship

LW and GNgMK conceptualised the study and completed the ethics application.

LW was the lead investigator for the study, completed the analysis and wrote the manuscript. All other authors completed data collection and provided feedback on the manuscript.

AG and TG are LW’s academic supervisors and provided support with development of this manuscript.

LW is the corresponding author.

## Data Sharing Statement

The anonymised data supporting the findings of this study are available from the corresponding author upon reasonable request.

## AI Use Statement

There has been no use of AI in the development of this study or manuscript.

## Ethics

The study was approved by the Health Research Authority (reference 23/HRA/4562). Further approval from a Research Ethics Committee was not required due to use of retrospective secondary data. Secondary data was managed and used as per GDPR guidelines.

