## Supplementary Table 1; Supplementary Figure 1 for "Mortality and diagnostic practice variation in interstitial lung disease admissions: insights from a multicentre UK cohort study"

**Supplementary Materials**

| ICD-10 Code | Diagnosis |
|---|---|
| B22.1 | HIV disease resulting in pneumocystosis |
| D86.0 | Sarcoidosis of lung |
| D86.2 | Sarcoidosis of lung and lymph node |
| J67.0 | Farmer's lung |
| J67.1 | Bagassosis |
| J67.2 | Bird Fancier's lung |
| J67.3 | Suberosis |
| J67.4 | Malt Worker's lung |
| J67.5 | Mushroom Worker's lung |
| J67.6 | Maple bark-stripper's lung |
| J67.7 | Air-conditioner and humidifier lung |
| J67.8 | Other hypersensitivity pneumonitis due to organic dusts |
| J67.9 | Hypersensitivity pneumonitis, unspecified |
| J70.2 | Acute drug-induced interstitial lung disorders |
| J70.3 | Chronic drug-induced interstitial lung disorders |
| J70.4 | Drug-induced interstitial lung disorders, unspecified |
| J84.1 | Other interstitial pulmonary diseases with fibrosis |
| J84.8 | Other specified interstitial pulmonary diseases |
| J84.9 | Interstitial pulmonary disease, unspecified |

**Supplementary Table 1:** ICD-10 codes with full diagnostic names used to identify admissions for inclusion within the study.

CONSORT

TRANSPARENT REPORTING of TRIALS

**CONSORT 2010 Flow Diagram**

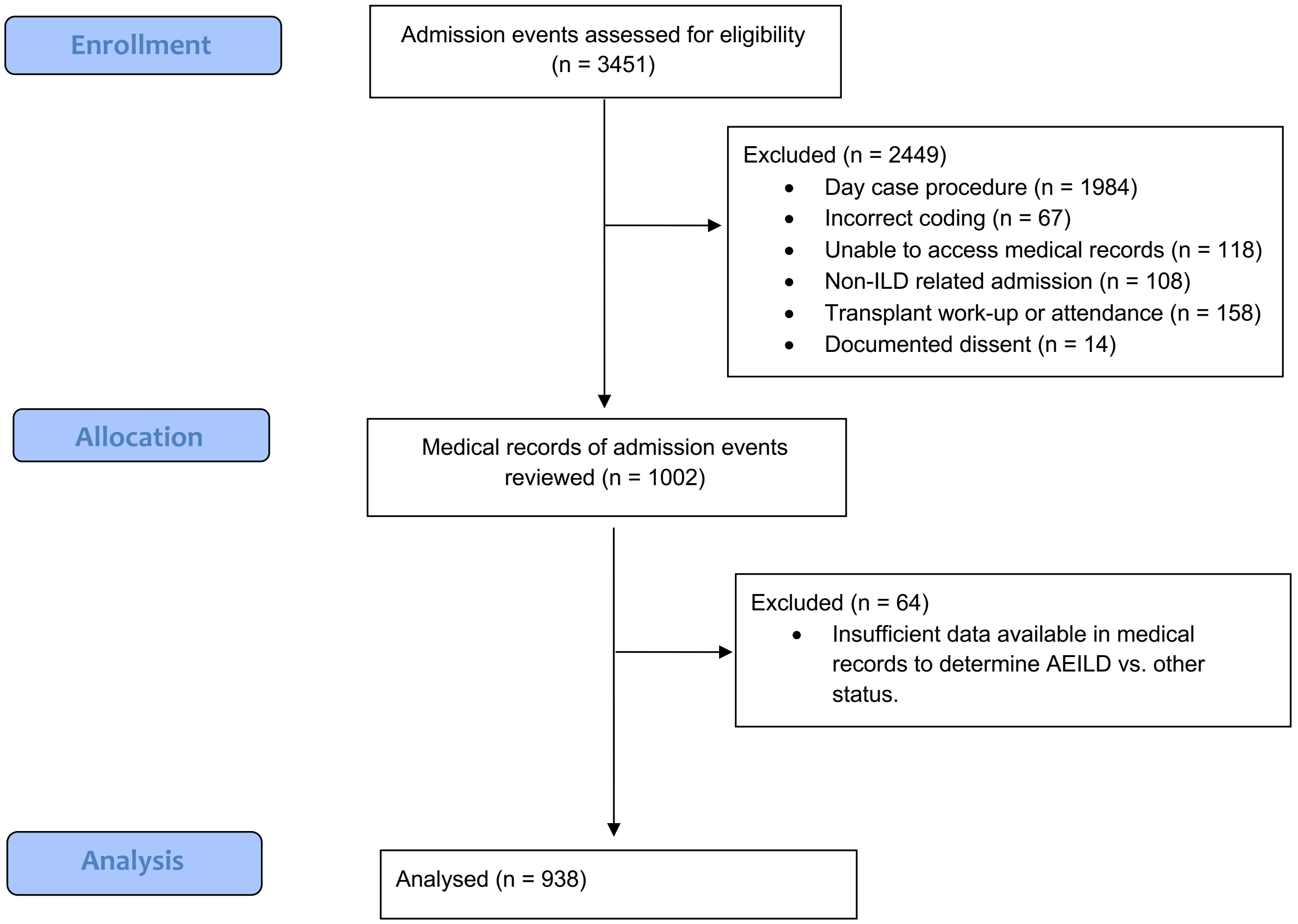


<u>**Supplementary Figure 1:**</u> CONSORT study flow diagram summarising records identified, reasons for exclusion and number of records included in analysis.

*Abbreviations: ILD – interstitial lung disease; AE-ILD - acute exacerbation of interstitial lung disease; ICD-10 – international classification of diseases version 10.*
